# Diagnosis and Prediction of Precursor States in Multiple Myeloma using Cell-free Chromatin

**DOI:** 10.1101/2025.10.31.25338878

**Authors:** Noa Omer Vilk, Vladimir Veinstein, Nadav Hermoni, Orly Ben-nun Shaul, Galina Pogrebijski, Esther Harpenas, Eyal Lebel, Marjorie Pick, Jenia Gutin, Moshe E. Gatt, Nir Friedman

**Affiliations:** The Lautenberg Center for Immunology and Cancer Research; Faculty of Medicine, The Hebrew University of Jerusalem, Israel; The Rachel and Selim Benin School of Computer Science and Engineering, The Hebrew University of Jerusalem, Israel; The Hebrew University Computational Medicine Center, The Hebrew University of Jerusalem, Israel; Department of Hematology, Hadassah Medical Center, Faculty of Medicine, The Hebrew University, Jerusalem, Israel; Senseera LTD, Jerusalem, Israel

**Author notes:** Equal contribution (Noa Omer Vilk, Vladimir Veinstein).

## Abstract

Multiple myeloma (MM) is a hematological malignancy that, in many cases, is preceded by monoclonal gammopathy of undetermined significance (MGUS) and smoldering multiple myeloma (SMM), pre-MM conditions with a lifelong risk of progression to symptomatic MM. Accurate diagnosis, prognostication, and adequate progress monitoring for these conditions require repeated invasive bone marrow biopsies. Additionally, current prognostic models for identifying higher-risk MGUS and SMM patients misclassify 20-30% of patients. Here, we apply Chromatin immunoprecipitation of cell-free chromatin (cfChIP-seq) from peripheral blood, which provides a genome-wide map of active promoters in the cells contributing to the circulating DNA, on a cohort of individuals with MGUS (n=65), SMM (n=27), patients with overt MM (n = 47), and healthy controls (n=82). We identified an increased representation of specific regions containing promoters specific to B-cells, plasma cells, and erythroblasts in MM blood samples. We used these regions to generate an MM score. This score is capable of diagnosing and staging the disease, and most importantly, identifying patients with a higher risk of 2-year progression (n = 18, p-value << 0.01). Our study highlights the potential of cfChIP-seq to contribute to differentiating plasma cell disease stages and monitoring progression non-invasively.

## Introduction

Multiple Myeloma (MM) is a complex and aggressive hematological malignancy characterized by the clonal proliferation of plasma cells within the bone marrow. This expansion disrupts normal bone marrow functions and leads to a range of clinical symptoms, including bone pain, fractures, anemia, hypercalcemia, and renal impairment^1^. Multiple myeloma (MM) is considered the final stage of plasma cell disorders (PCDs), which begins with monoclonal gammopathy of undetermined significance (MGUS) and progresses through smoldering multiple myeloma (SMM) before transforming into symptomatic, active disease. This progression is highly variable, with the majority of patients remaining in the early stages for years while others rapidly develop active disease.^2,3^

Diagnosis and staging of MM are essential for effective treatment. International Myeloma Working Group (IMWG) criteria for MM diagnosis require at least 10% clonal plasma cells in the bone marrow or a biopsy-proven plasmacytoma, along with myeloma-defining events (MDEs).^4^

Despite advances in understanding the biology of MM, several significant challenges remain in predicting disease behavior and managing asymptomatic stages. For patients with asymptomatic MGUS or SMM, the primary concern is the risk of progression to overt MM. The clinical dilemma is identifying the optimal timing for therapeutic intervention, as treating too early may lead to unnecessary toxicity, while delaying treatment may allow irreversible organ damage. However, predicting whether and when a progression will occur is an unmet need.

Current practices for diagnosis and prognosis in the spectrum of plasma cell disorders (PCDs) include the IMWG assessment for SMM, and the Mayo Clinic score for MGUS^5–7^. The IMWG assessment relies on bone marrow biopsies, which are invasive and painful, making repeated assessments difficult. Additionally, biopsies may miss disease areas due to uneven distribution of plasma cells, leading to inaccurate assessments. Moreover, currently available scoring systems for SMM do not have high predictive value – 20-40% of low- to intermediate-risk patients progress within 2 years, while 30-40% of high-risk patients do not. This highlights the need for scoring systems with increased accuracy, preferably using non-invasive and easily repeatable techniques.

Suggestions for such newer approaches span circulating tumor cells, exosomes, proteins, miRNA, and more^8,9^. Previous studies on cell-free DNA (cfDNA) based on mutations and copy number aberrations have shown that plasma cells contribute varying amounts to cfDNA, with levels rising with disease progression.^10^

Recently, we developed chromatin immunoprecipitation and sequencing of cfDNA (cfChIP-seq) to survey the epigenetic state of cells contributing to the circulating DNA pool ^11^. Briefly, this assay exploits the structure of circulating DNA, mainly in the form of intact nucleosomes that retain histone modifications from the cell of origin. Through immunoprecipitation, we enrich for cell-free nucleosomes that carry specific histone modifications of active promoters (Histone 3 Lysine 4 tri-methylation, H3K4me3). Sequencing the DNA from these nucleosomes enables us to map the genomic regions associated with active promoters.

Here, we examine cfChIP-seq as a noninvasive tool for diagnosing and monitoring progression in PCDs. We collected blood samples from patients with overt MM, individuals with MGUS and SMM, and from healthy subjects. We hypothesized that applying cfChIP-seq to these samples would identify genomic regions with differential signals distinguishing between healthy individuals, those in pre-MM stages, and patients with overt MM, and that higher levels of these regions would identify patients at high risk of progressing from pre-malignant stages to overt MM.

## Results

### Study participants

Blood samples were collected from 82 self-reported healthy individuals, 65 individuals with MGUS, 27 individuals with SMM, and 47 patients with overt MM (Fig. 1A). Blood samples of patients with overt MM were taken prior to treatment. The characteristics of study participants are summarized in Table 1 and detailed in Table S1.

**Figure 1:**
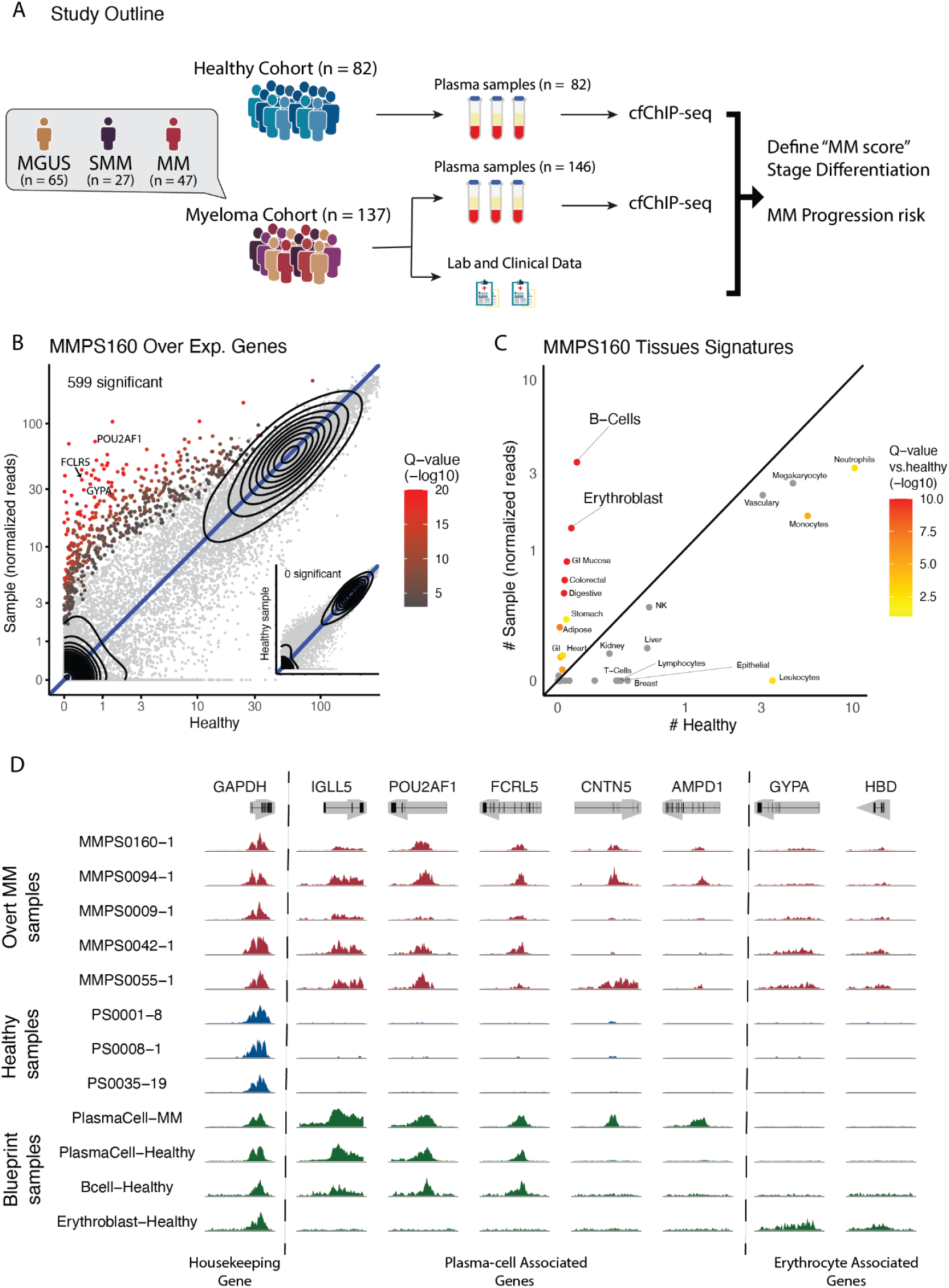
Cell-free chromatin in plasma of MM patients. A. Study outline - Samples were collected from healthy and myeloma patients at different stages. All samples were processed using the cfChIP-seq protocol, and relevant clinical data were collected. **B. Genes with significantly elevated levels in a sample** - When comparing the genes of a patient with overt MM to those of healthy controls, we can see that many genes are significantly elevateed, indicating that the cfDNA composition significantly higher from that of a healthy individual. **C. Tissue signatures in a sample** - Using published ChIP-seq data to define cell type/tissue-specific signatures as promoters with high signal only in one cell type, we can see that these overrepresented regions are areas related to B-cells and Erythoblasts. **D. Genome browser view of cfChIP-seq signal in plasma cell and Erythroblast known genes.** The top tracks (red) are cfChIP-seq signals from five myeloma patients. The middle tracks (blue) are cfChIP-seq signals from three healthy donors, and the lower tracks (green) are Blueprint samples run on sorted cell populations.<u>(Adams et al. 2012)</u> The dashed line separates housekeeping genes, plasma-cell-associated genes, and erythrocyte-associated genes.

**Table 1:** Cohort characteristics.

|  | Healthy | MGUS | SMM | overt MM | p |
| --- | --- | --- | --- | --- | --- |
| Age | 31.80<br>[28.8–34.7] | 68.45<br>[66.0–70.9] | 69.78<br>[65.0–74.5] | 66.72<br>[63.1–70.4] | <0.001 |
| Gender |  |  |  |  |  |
| F | 38 (50.7%) | 22 (31.4%) | 13 (44.8%) | 22 (46.8%) | 0.115 |
| M | 37 (49.3%) | 48 (68.6%) | 16 (55.2%) | 25 (53.2%) |  |
| Immunoglobulin Type |  |  |  |  |  |
| FLC |  | 15 (21.4%) | 5 (17.2%) | 13 (27.7%) |  |
| IgA |  | 8 (11.4%) | 7 (24.1%) | 0 |  |
| IgG |  | 26 (37.1%) | 16 (55.2%) | 6 (12.8%) |  |
| IgM |  | 2 (2.9%) | 1 (3.4%) | 0 |  |
| BM infiltration (%) |  | 0.03<br>[0.02–0.04] | 0.19<br>[0.11–0.28] | 0.41<br>[0.27–0.56] | <0.001 |
| Albumin |  | 29.55<br>[24.7–34.4] | 20.67<br>[13.2–28.1] | 24.00<br>[18.1–29.9] | 0.1 |
| B2-Microglobulin |  | 3.44<br>[2.02–4.86] | 2.89<br>[2.24–3.54] | 5.87<br>[3.97–7.76] | 0.0332 |
| Creatinine |  | 64.13<br>[51.9–76.4] | 42.01<br>[23.9–60.1] | 68.51<br>[35.0–102.0] | 0.329 |
| Erythroblast-enriched subscore |  | 139.5<br>[87–192] | 121.1<br>[48–194] | 533.8<br>[321–746] | <0.001 |
| Plasma-cell enriched subscore |  | 37.7<br>[28–47] | 46.8<br>[28–66] | 354.7<br>[213–497] | <0.001 |
| cfChIP MM score |  | 177.3<br>[120–234] | 167.9<br>[87–249] | 888.6<br>[627–1151] | <0.001 |
| ISS Score (MM only) |  |  |  | 1.94<br>[1.67–2.21] |  |

Plasma samples from this cohort were analyzed using cfChIP-seq targeting nucleosomes marked by H3K4me3, a mark of poised and active promoters (Table S2, Methods, Fig. S1A). Only samples that passed our quality control thresholds, based on the number of unique and informative reads and background rate, were included in the final cohort. ^11^

### Distinct cfChIP-seq signals in patients with MM originating from plasma cells and erythrocyte progenitors

Examining samples from patients with overt MM revealed signals that deviate from the healthy reference (Fig. S2A). For example, a sample from patient MMPS160 shows 559 gene promoters whose level is significantly elevated compared to a healthy baseline (Fig. 1B, Methods). Many of these genes (e.g., POU2AF1, FCRL5, GYPA) are rarely found in samples from healthy controls. Examining the cell types contributing to the circulation, we observed a significant increase in B cells and erythroblasts (Fig. 1C), suggesting increased cell death or turnover of B-cell and erythrocyte lineages. We observe additional signatures with abnormally high values. Similar patterns are observed in samples from other patients with overt MM (Fig. S2B, S2C).

Genes with significantly elevated levels that were common to these samples included erythroblast-specific genes such as Glycoporin A (GYPA) and Hemoglobin delta (HBD); B-cell and plasma cell specific genes, such as, Immunoglobulin lambda like polypetide 5 (IGLL5)^12^; and genes associated with malignant plasma cells in MM such as FC receptor like 5 (FCRL5), Lymphocyte Transmembrane Adaptor 1 (LAX1), and POU Class 2 Homeobox Associating Factor 1 (POU2AF1)^13–15^ (Fig 1D). The levels of these promoters in healthy individuals were below detection levels but significantly higher in samples from patients with overt MM.

To validate these findings, we compared cfChIP-seq signals to previously published H3K4me3 ChIP-seq profiles from the Blueprint Epigenome Project. Viewing these significantly elevated genes, such as *FCRL5*, *POU2AF1*, *GYPA*, and *HBD*, we observe that they are also elevated in Blueprint reference samples from plasma cells and erythroblasts (Fig. 1D). These findings suggest that cfChIP-seq can detect abnormal signals in samples from patients with overt MM. In particular, these signals include disease-related active genes originating from malignant plasma cells (e.g., FCRL5 and LAX1) and disease-induced damage to normal hematopoiesis (e.g., erythrocyte lineage).

### Defining a cfChIP-seq-based MM score

We compared the cfChIP-seq profile in a training set that included samples from patients with overt MM (n = 30 selected randomly from 47 overt MM samples; Table S1) and from healthy controls (n = 82). We devised a likelihood-based test that accounts for the variation in disease contribution within each sample (Methods, Supplementary Note). We identified 302 genomic regions that exhibited significantly higher cfChIP-seq signal levels in samples from patients with overt MM (p < 0.01), with at least a 3-fold increase in levels compared to samples of healthy controls (Fig. 2A). The identified regions overlap with promoters of 62 annotated RefSeq genes, which are enriched for genes associated with erythrocytes (26 genes, *p* < 10^-7^) and plasma cells (17 genes, *p* < 0.02).^16^

**Figure 2:**
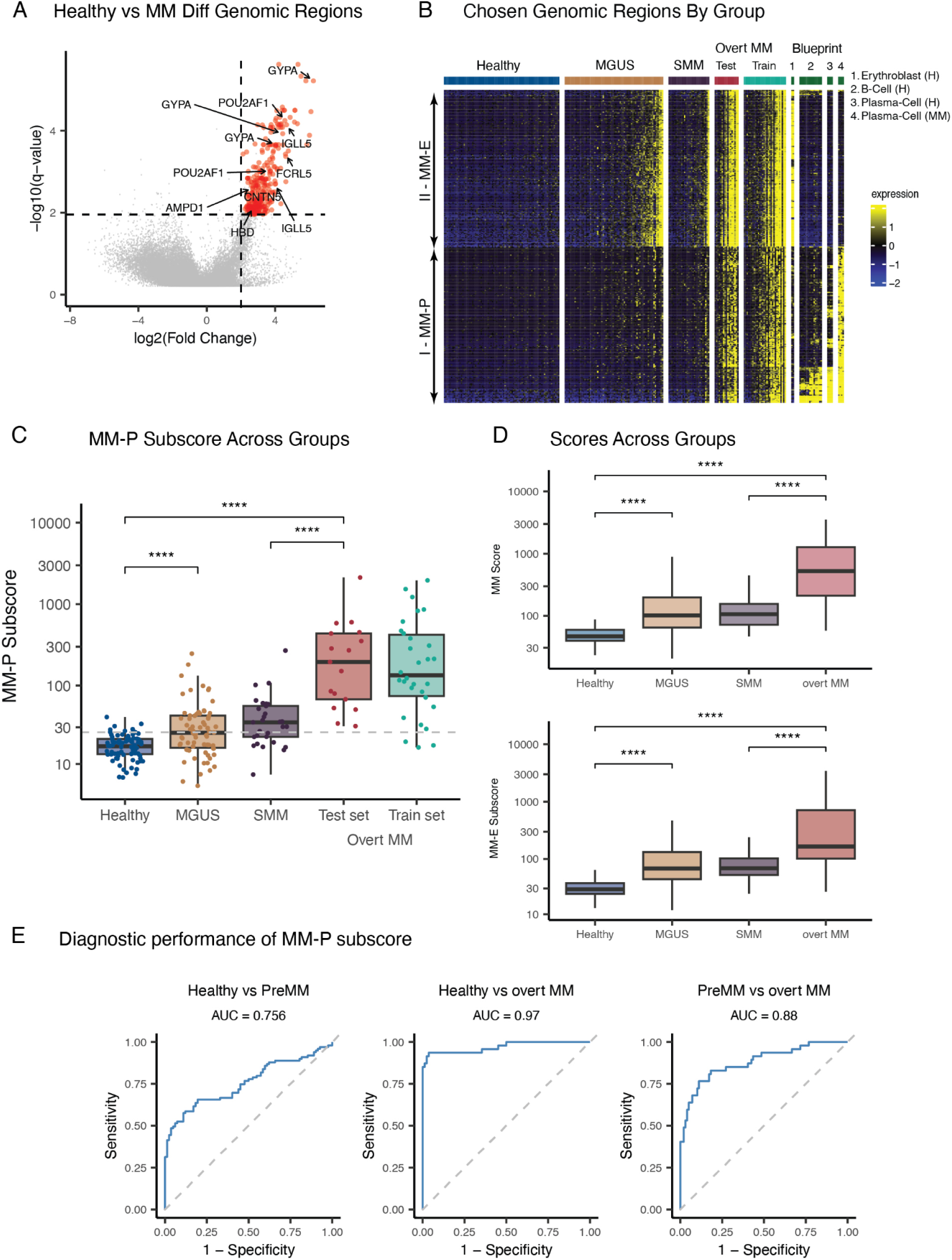
Identification and performance of MM-associated genomic regions **A. Differential Genomic Regions Analysis** - Volcano plot showing differential signal between overt MM and healthy samples. Red points represent regions with significant signal (adjusted p < 0.01, FC > 3). **B. Heatmap of Selected Genomic Regions** - Heatmap of 302 selected genomic regions across samples. Samples are grouped by clinical category: Healthy, MGUS, SMM, and overt MM (divided into train and test). Reference samples from the Blueprint project are included for comparison, representing healthy erythroblasts, plasma cells, B cells, and MM plasma cells. Signal intensity is color-coded (yellow = high, blue = low). **C. MM-P Subscore by Group** - Distribution of Plasma cell-enriched (MM-P) subscore across clinical groups. Subscore increases with disease progression. Statistical significance assessed via Wilcoxon tests (**** p < 0.0001). **D.** Top - Distribution of MM score across clinical groups, combining both subscores. The score increases along the disease spectrum. Bottom - Distribution of Erythroblast-enriched (MM-E) subscore across clinical groups. This subscore also increases with disease stages but displays higher discriminative power between healthy and early-stage disease **E. ROC Curves for Group Classification** - ROC curves evaluating the ability of the MM-P subscore to distinguish clinical groups. AUC values are shown for each pairwise comparison.

Examining the cfChIP-seq signal across the selected regions, we observed consistently elevated levels in overt MM samples—both in the test cohort and in the overt MM samples from the training set (Fig. 2B). In contrast, pre-MM samples showed overall lower signal, although some signal was still detectable (Fig. 2B). To quantify this observation, we summarized the signal across all regions to derive an MM score for each patient, reflecting disease activity and progression potential (Fig. 2C).

### Distinct Gene Clusters as MM subscores

To further understand the MM signal and its biological significance, we clustered the regions (Methods). The resulting clustering matrix divides into two equal-sized subscores (Fig. 2B, S3A). Regions in one subscore (II) were tightly intercorrelated, while the ones in the other cluster (I) showed lower internal correlation. Examining the genes in each cluster (Table S3), we observed that genes in subscore I significantly overlap with plasma cell-specific genes (11 genes, p < 0.002), and genes in subscore II significantly overlap with erythrocyte-specific genes (26 genes, p < 10^-13^). Notably, all three scores remained relatively consistent across age groups (Fig. S3B), indicating that score variation is not driven by patient age.

Evaluating the two subscores across ChIP-seq profiles of sorted immune cell populations^17^ further supported this relationship. Subscore I is high in plasma cells, particularly in plasma cells from patients with overt MM, and to a lesser degree in B cells, while subscore II is high in erythroblasts (Fig. 2B, S3C). We will thus refer to these two groups as Plasma cell-enriched (MM-P subscore) and Erythroblast-enriched subscores (MM-E subscore), respectively. We observed a strong correlation between the MM-P and MM-E subscores across samples (Fig. S3D), suggesting that both subscores capture overlapping yet distinct aspects of disease biology and progression.

### cfChIP-seq MM Score Increases With Disease Stage

The MM score and its subscores, MM-P and MM-E, all showed significant differences between healthy individuals and each disease stage—pre-MM and overt MM—suggesting that pre-MM stages already exhibit myeloma-associated signals (Fig. 2C, 2D; p < 0.01). Interestingly, among them, MM-E showed the strongest separation between healthy and pre-MM samples (AUC = 0.88), followed by MM-P (AUC = 0.76) (Fig. 2E, S3E).

### MM Score Stratifies Risk For Disease Progression

We followed the clinical course of individuals with pre-MM conditions including those with at least one follow-up visit (n = 72) We identified two types of disease progression events: clinical progression, which involves the escalation of diagnosis to overt multiple myeloma (MM) and the initiation of treatment, and biochemical progression, where a patient’s laboratory work deteriorates according to parameters defined by the IMWG criteria^7^ (Methods). The date of treatment start marked clinical progression, whereas the date of the qualifying laboratory result marked biochemical progression.

We first assessed the association between 24-month progression and the three scores: the global MM score and the MM-P and MM-E subscores. ROC analysis (Fig. S4A) showed that the MM-P score was the most informative. For clinical progression, MM-P achieved an AUC of 0.67, compared with 0.60 for the MM score and 0.57 for MM-E. For biochemical progression, MM-P reached an AUC of 0.71, versus 0.64 and 0.61, respectively. MM-P offered the strongest discrimination, and subsequent analyses focused on this subscore.

To test whether MM-P also stratifies time to progression, we defined high-risk individuals as those above the median MM-P value within their diagnostic group (MGUS or SMM). Kaplan–Meier curves revealed a significant difference in biochemical progression-free survival between high- and low-risk groups (Fig. 3A). A similar, but borderline, separation was observed for clinical progression, probably owing to the smaller number of clinical events (Fig. 3B).

**Figure 3.**
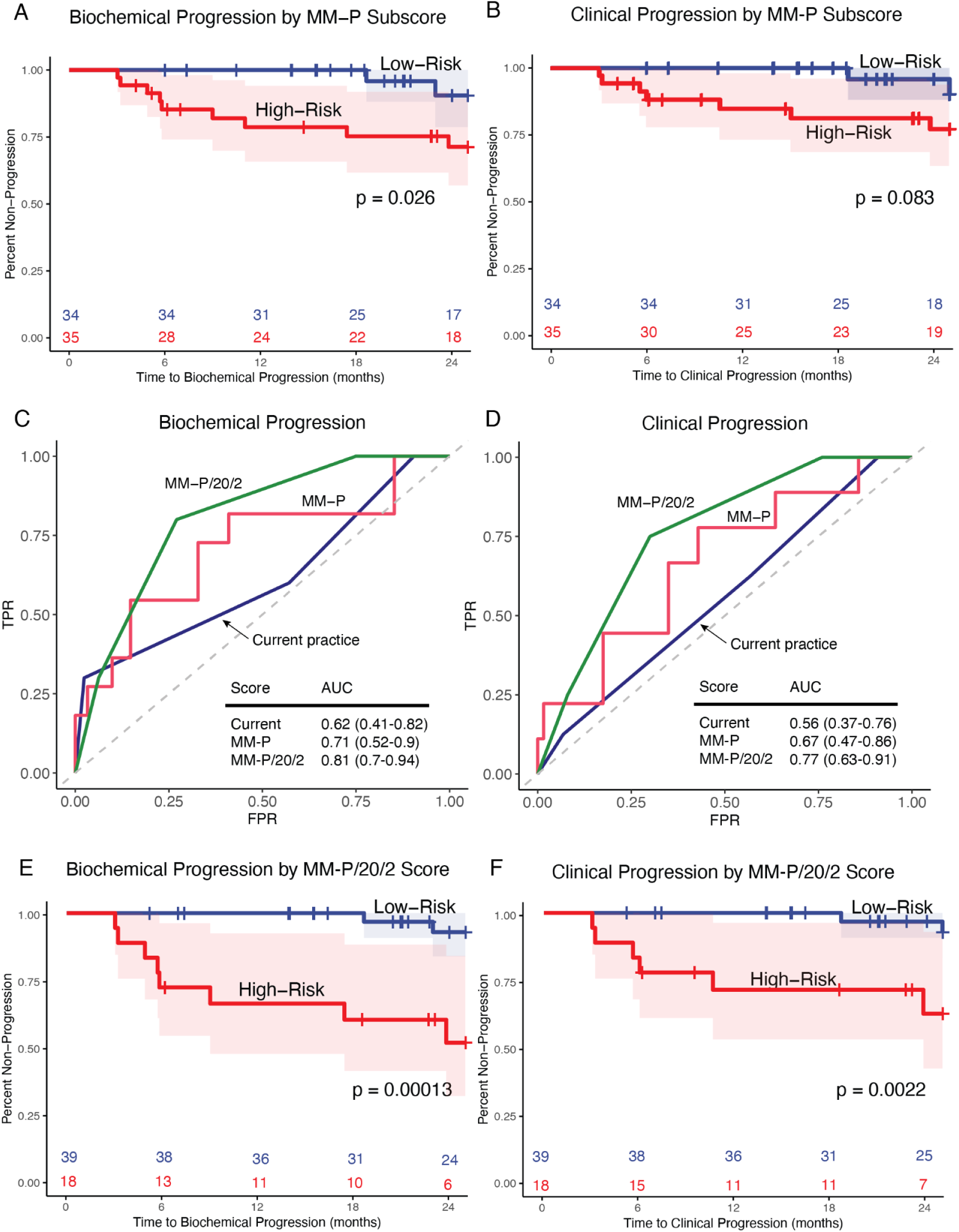
cfChIP-derived MM-P score improves risk stratification for progression in individuals with pre-MM. **A. Time to Biochemical Progression by MM-P Subscore** – Kaplan–Meier analysis comparing high-risk and low-risk individuals based on MM-P subscore. High-risk individuals show significantly faster biochemical progression (log-rank p = 0.026). **B. Time to Clinical Progression by MM-P Subscore** – Kaplan–Meier curve for clinical progression using the same MM-P stratification. A trend toward faster progression in the high-risk group is observed (log-rank p = 0.083). **C. ROC Curve for Biochemical Progression (24 months)** – ROC analysis comparing: (i) standard clinical risk models (Mayo score for MGUS and IMWG 20/2/20 for SMM; orange, AUC = 0.61), (ii) the MM-P subscore alone (green, AUC = 0.71), and (iii) the combined MM-P/20/2 score integrating MM-P with clinical variables (blue, AUC = 0.81). **D. ROC Curve for Clinical Progression (24 months)** – ROC analysis for predicting clinical progression using MM-P/20/2 (AUC = 0.76), MM-P alone (AUC = 0.67), and clinical risk scores (AUC = 0.56). **E. Time to Biochemical Progression by MM-P/20/2 Score** – Kaplan–Meier curve stratified by the integrated MM-P/20/2 score, combining MM-P with IMWG 20/2/20 criteria. The score robustly distinguishes high- and low-risk individuals (log-rank p = 0.00084). **F. Time to Clinical Progression by MM-P/20/2 Score** – Kaplan–Meier analysis of clinical progression stratified by MM-P/20/2 score (log-rank p = 0.0081), showing improved separation compared to MM-P alone.

We note that the risk for progression was not associated with MGUS vs. SMM distinction, and in both groups, the individuals with “low risk” samples showed less progression (Fig. S4A). Similarly, other possible confounders (age, sex, Hemoglobin (g/dL)) do not explain the difference in time to progression (Fig. S4B).

### cfChIP-seq MM Score Correlates with Common Prediction Scores

Current clinical guidelines for monitoring individuals with pre-MM rely on risk-stratification models to assess the likelihood of disease progression. For MGUS patients, the Mayo Clinic model^5^ identifies three major risk factors: serum M-protein concentration >1.5 g/dL, non-IgG isotype (e.g., IgA or IgM), and an abnormal serum free light chain (FLC) ratio (<0.26 or >1.65). Each risk factor is assigned one point, categorizing patients into low (0 points), intermediate (1 point), or high-risk (2-3 points) groups. This stratification helps determine appropriate monitoring intervals and management strategies. For individuals with SMM, the International Myeloma Working Group (IMWG) employs the “2/20/20” model^7^, which includes serum M-protein level >2 g/dL, bone marrow plasma cell infiltration >20%, and FLC ratio >20. Patients are classified as low risk (0 factors), intermediate risk (1 factor), or high risk (2-3 factors).

We assigned a score to each sample using the conventional clinical risk schemes (IMWG for SMM and the Mayo score for MGUS). ROC analysis showed that the cfChIP MM-P sub-score consistently outperformed the clinical score. For clinical progression within 24 months, MM-P reached an AUC of 0.67, whereas the IMWG/Mayo score achieved 0.56. For biochemical progression, MM-P achieved an AUC of 0.71 compared with 0.61 for the clinical score (Fig. S4C).

### Integrating Plasma Cell-Enriched Subscore with Clinical Criteria Enhances Risk Stratification

To explore whether integrating traditional clinical metrics with our cfChIP-seq derived MM score could further improve risk prediction, we generated a modified version of the IMWG “2/20/20” model. In this adapted model, we replaced the bone marrow plasma cell infiltration criterion (>20%), which requires an invasive biopsy, with the cfChIP-seq MM-P subscore, using a threshold corresponding to the 50th percentile in our dataset (as explained above). A patient is considered high risk if they meet two or more of the following: MM-P > median, FLC ratio > 20, M-protein >2 g/dL. This hybrid MM-P/20/2 score improved discrimination, raising the AUC to 0.81 for biochemical progression and 0.76 for clinical progression (Fig. S4C). Kaplan–Meier analyses confirmed robust stratification between the high- and low-risk groups (log-rank p = 0.00001 and 0.002, for biochemical progression and clinical progression, respectively; Fig. 3E, F).

We next evaluated each component of the MM-P/20/2 score individually using univariate Cox regression (Fig. S4D). FLC ratio ≥20 was significantly associated with both biochemical and clinical progression (p = 0.01 and 0.01, respectively), while M-protein ≥2 g/dL did not show significant associations (p = 0.53 and 0.84). The MM-P subscore, using a median threshold, showed a nominal association with biochemical progression (p = 0.08) and a non-significant trend for clinical progression (p = 0.17).

## Discussion

We set out to understand circulating free DNA (cfDNA) in patients with plasma cell disorders (PCDs), specifically how it can inform us about the disease state and severity. Current diagnostic and monitoring approaches for MM rely heavily on invasive bone marrow biopsies, blood tests measuring serum protein levels, and imaging studies. However, these approaches have notable limitations: bone marrow biopsies are invasive, limiting their frequency and feasibility for ongoing monitoring; serum protein-based blood tests vary in sensitivity and specificity, particularly in early or pre-symptomatic disease stages; and imaging studies may not capture subtle or systemic disease changes. These limitations underscore the need for a sensitive and comprehensive method to detect disease-related signals at different stages.

To address these limitations, we applied cfChIP-seq, a non-invasive technique that analyzes circulating chromatin fragments in plasma. We identified clear cfChIP-seq signals in patients with overt MM originating from abnormal plasma cells and additional cellular sources. Notably, these signals were elevated even in pre-symptomatic stages (MGUS and SMM) and intensified with disease progression toward symptomatic MM.

The cfChIP-seq-derived score integrated two components: a plasma cell-associated score and an erythroblast-associated score. The plasma cell component includes a mix of genes that are shared across malignant and non-malignant plasma cells, genes uniquely active in malignant cells, and genes expressed in B cells, capturing gene expression patterns associated with the MM disease process. These genes may serve as early indicators of advancing pathology. The erythroblast-derived signals indicate increased erythrocyte production conditions (Sadeh et al. 2021; Moss et al. 2023), which occur even before clinical anemia is apparent in symptomatic MM.

Although the combined score effectively distinguished healthy individuals from patients with plasma cell disorders, it did not clearly separate MGUS from SMM cases. This limitation aligns with the clinical challenge of distinguishing between MGUS and SMM based on an arbitrary cutoff of 10% plasma cells in the bone marrow. Since these precursor states may remain stable for years before progressing, we specifically used the plasma cell subscore for progression analyses. This subscore allowed us to focus more precisely on disease processes during pre-MM stages. When evaluating 24-month progression outcomes, we observed that individuals with a higher plasma cell–enriched subscore had a significantly increased risk of progressing to overt MM.

Extending this concept, we integrated the plasma-cell subscore with two available serum markers from the clinically used IMWG 2/20/20 scoring system, M-protein and FLC ratio, creating the MM-P/20/2 score. By substituting the invasive marrow-infiltration criterion of the original IMWG 20/2/20 model with our cfChIP score, the hybrid score preserved the “two-of-three” rule while eliminating the need for invasive biopsies. This combination outperformed each component alone as well as the original 20/2/20 score, increasing the AUC to 0.81 for biochemical progression and 0.76 for clinical progression, and it stratified time to both biochemical and clinical progression with log-rank p-values of 0.0008 and 0.008, respectively. This suggests that cfChIP-seq can be seamlessly combined with routine laboratory tests to create a fully non-invasive yet highly informative tool for early risk assessment in individuals with pre-MM. Adoption of this approach could streamline monitoring schedules, prioritize high-risk patients for closer follow-up or early intervention studies, and ultimately reduce the biopsy burden on this already vulnerable population.

While our current study focused on stratifying pre-MM patients, similar approaches could be valuable for assessing the risk of relapse post-treatment. Identifying common features across patient samples is essential, but further research into distinct disease subtypes and their therapeutic implications remains an important next step.

In conclusion, our method uniquely captures comprehensive genomic signals related to MM in a non-invasive manner, detecting clinically relevant cell populations beyond malignant plasma cells. These results complement approaches based on DNA methylation^22^, which achieve a similar task by detecting plasma-cell lineage markers and aberrant patterns of DNA modifications. Together, both methods present exciting advances toward non-invasive tools for managing pre-MM patient populations.

## Supporting information

Clinical Summary

Supplemental Table 3 - Genes in Score

Supplemental Table 1 - Clinical data

## Data Availability

Genome browser tracks and tables of gene levels in all samples are available in the Zenodo repository: https://doi.org/10.5281/zenodo.1563478. (Reviewers’ accessible link https://tinyurl.com/mrnnbrjj)

## Acknowledgements

We thank Gavriel Fialkoff, Ronen Sadeh, Shoshana Revel-Vilk, and members of the Friedman lab for comments and suggestions. This work was supported in part by an ERC grant (#101019560) and Rosetrees Foundation Project Grant (#PGL22/100002).

## Conflict of Interests

J.G. and N.F. are founders and shareholders of Senseera LTD. All other authors declare no potential conflicts of interest.

## Methods

### Sample cohort

Blood samples were collected from patients with overt MM, individuals with SMM and MGUS (pre-MM), followed at the Department of Hematology, Hadassah Medical Center. Samples from patients with MM were taken before initiation of therapy. All study participants signed informed consent forms before sample collection following a study protocol approved by the Hadassah Medical Center Institutional Review Board (IRB) committee. Healthy samples were collected from self-declared healthy individuals at the Hebrew University.

Blood samples were processed following previously described methods ^11^. In brief, plasma was separated from whole blood by two centrifugations, and cell-free chromatin was isolated using immunoprecipitation targeting histone modifications associated with active promoters. The resulting DNA was purified and prepared for sequencing to generate genome-wide profiles of promoter activity.

Clinical assessments and laboratory data of patients with overt MM, and individuals with SMM and MGUS (pre-MM) were collected from electronic health records.

### Control group

We used the healthy cohort to create a reference profile representing the expected cfChIP-seq profile of a healthy individual (mean and variance for each gene). This reference profile served as a baseline to identify genomic regions in other samples that deviate from the healthy profile, indicating potential contributions from non-healthy or disease-associated tissues. Comparing individuals from different age groups showed little differences (Fig. S1B), suggesting that this baseline is stable with age.

### MM score

#### Training sets

To identify the differential regions between healthy and MM samples, we utilized the samples of patients with overt MM and samples from healthy controls. The patient samples were randomly divided into a 70-30 split for training and testing. The 70% group was used to train the MM score, while the remaining 30%, along with the SMM and MGUS samples, formed the test set. All control samples were used to refine the score.

#### Window preselection

We used a catalog of genomic windows covering the human genome as previously described ^11^. This catalog defines ∼1.6M windows covering the non-excluded regions of the genome. Window size ranges between 200bp and 5,000bp, based on the levels of H3K4me3 in the Roadmap Epigenetics samples^7,18^.

We implemented a preselection criteria method to eliminate genomic windows with high signal in healthy controls, high variability, or extreme fold changes between male and female samples. First, we calculated each genomic window’s mean and standard deviation using the healthy control sample data. To assess the stability of each genomic window, we used the coefficient of variation (CV), the ratio of the standard deviation to the mean, providing a normalized measure of variability. Next, we split the data from healthy controls by sex, calculating the mean values for male and female samples separately, and computed male-to-female fold-change to ensure that neither group exhibited extreme biases.

We retained windows with a male-to-female fold change below 1.5, focusing on regions with minimal sex-related differences. Additional filters included a CV less than one and a window mean below a threshold of 1 normalized count in samples from healthy controls. Windows that met these criteria (96% of all windows) were retained for further analysis. We further pruned the potential genomic windows to ones with non-negligible signal levels in the MM training samples (mean > 1), resulting in 40,883 potential windows.

#### Statistical analysis

We developed a statistical test that assumes that cfDNA in a sample from a patient with disease is a weighted combination of cfDNA from the healthy population and some contribution of disease tissue. The model assumes that the relative contribution of disease tissue in each sample varies, and estimates this quantity from the data (Supplemental Note). The test then compares a null hypothesis where there is no difference between the distribution of a window in both sources and the alternative hypothesis where there might be differences. We used a variation of the likelihood ratio test (LRT) to evaluate the p-value of the null hypothesis for each window (Supplemental Note).

The resulting p-values were adjusted for multiple comparisons using false discovery rate (FDR) ^19^. Then, we computed fold-change (FC) from the signal that remained after removing noise and other confounders. FC reflects only the inferred contributions from cell types other than neutrophils and megakaryocytes. Windows with *q* < 0.01 and *FC* < 3 were considered significantly high in MM.

Using the genomic windows selected in the previous step, we defined an MM score by summing their values to create a cumulative MM score for each patient.

### Subscores

To investigate gene correlation patterns, we generated a correlation matrix from the cfChIP-seq levels using Pearson’s correlation, which effectively captures linear relationships between the chosen regions. We then converted the correlation matrix into a distance matrix. Using hierarchical clustering, we constructed a dendrogram to identify groups of genes with similar level profiles (Supplementary code).

We divided the dendrogram into two clusters, effectively categorizing genes with similar cfChIP signal profiles. These clusters offer a more organized view of gene interactions. Each cluster was used to define a separate subscore (each containing 151 different genomic windows).

### Time-to-Progression Analysis

To define high-risk pre-malignant patients, who are more likely to progress to a malignant stage, we used a threshold based on the 50th percentile (median) of the MM score in samples from individuals with MGUS and SMM, separately. Using this threshold, we classified samples with scores above the threshold as “high risk” and those below the threshold as “low risk.”

Progression time was defined as follows:

- Clinical progression - initiation of treatment
- Biochemical progression - meeting one of the following criteria: M-protein increase > 0.5 g/dL, involved FLC increase > 25% with absolute FLC level > 100 mg/L.
- No progression—The individual had at least one follow-up evaluation after the initial sample, but there was no evidence of progression.

We performed a Kaplan–Meier (KM) survival analysis to evaluate time to progression, using the **log-rank test** to assess statistical significance between high- and low-risk groups. Risk groups were defined separately for MGUS and SMM patients based on the median value of the specified score within each group. Time to biochemical or clinical progression was calculated in months from diagnosis to event or censoring, capped at 24 months. The analysis was implemented in R (v. 4.3.2) using the survival (v. 3.7-0) and survminer (v. 0.4.9) R packages.

## Code availability

All script files used in this manuscript’s analysis will be available upon request. R code for processing cfChIP-seq data is available at https://github.com/nirfriedman/cfChIP-seq git, and set-up files for analysis are available in the Zenodo repository: https://doi.org/10.5281/zenodo.3967253.

## Supplementary Figures

**Figure S1:**
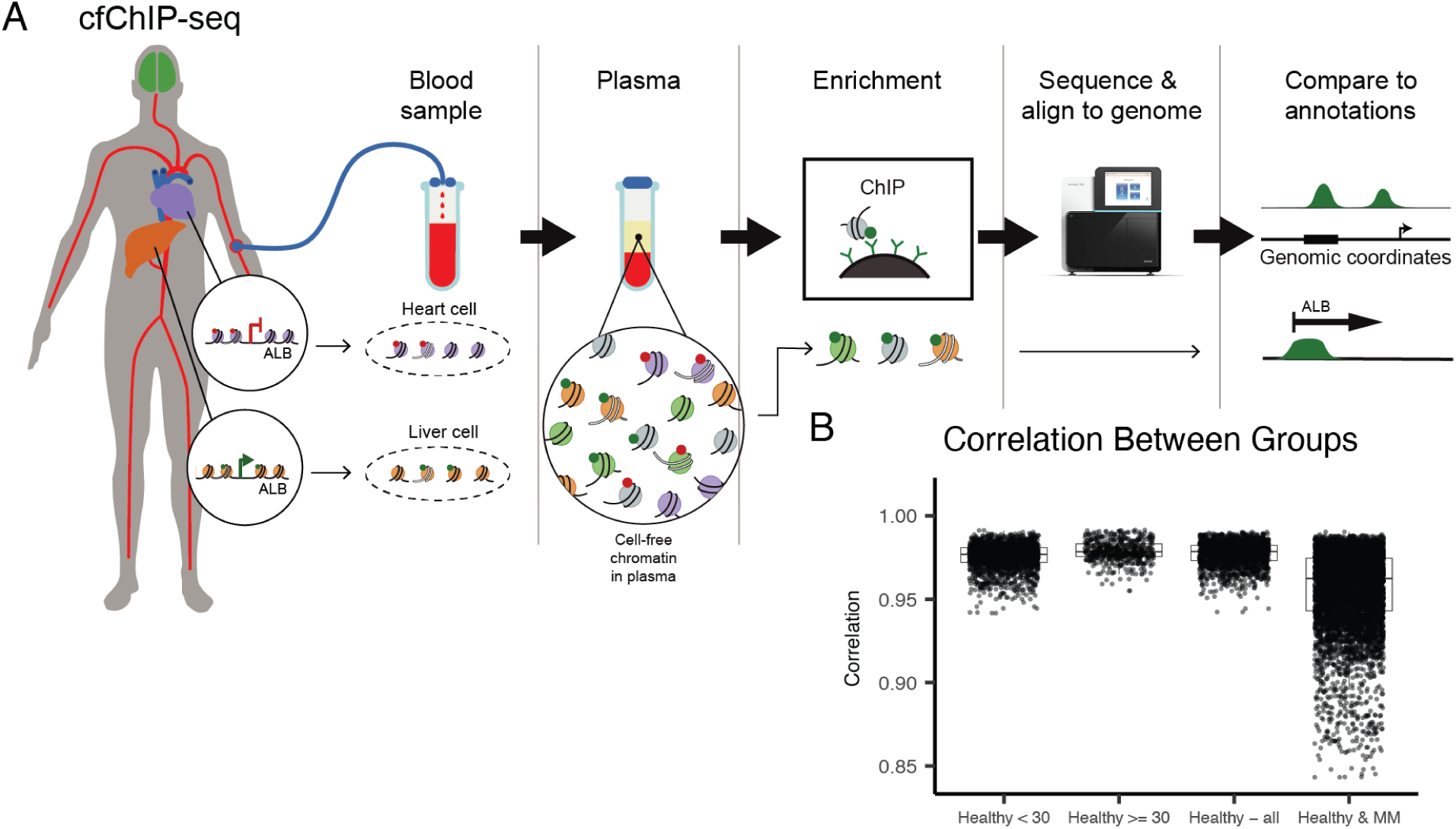
cfChIP-seq workflow and characterization of signal distribution across clinical stages A. cfChIP-seq Workflow. – Schematic overview of the cfChIP-seq protocol. Plasma is isolated from whole blood, and cell-free chromatin is enriched using immunoprecipitation of H3K4me3-marked nucleosomes. Enriched fragments are sequenced and aligned to the genome for downstream analysis. **B. Global Correlation of cfChIP-seq Signal** – Correlation of genome-wide cfChIP-seq signal across healthy samples (stratified by age) and patients with MM.

**Figure S2:**
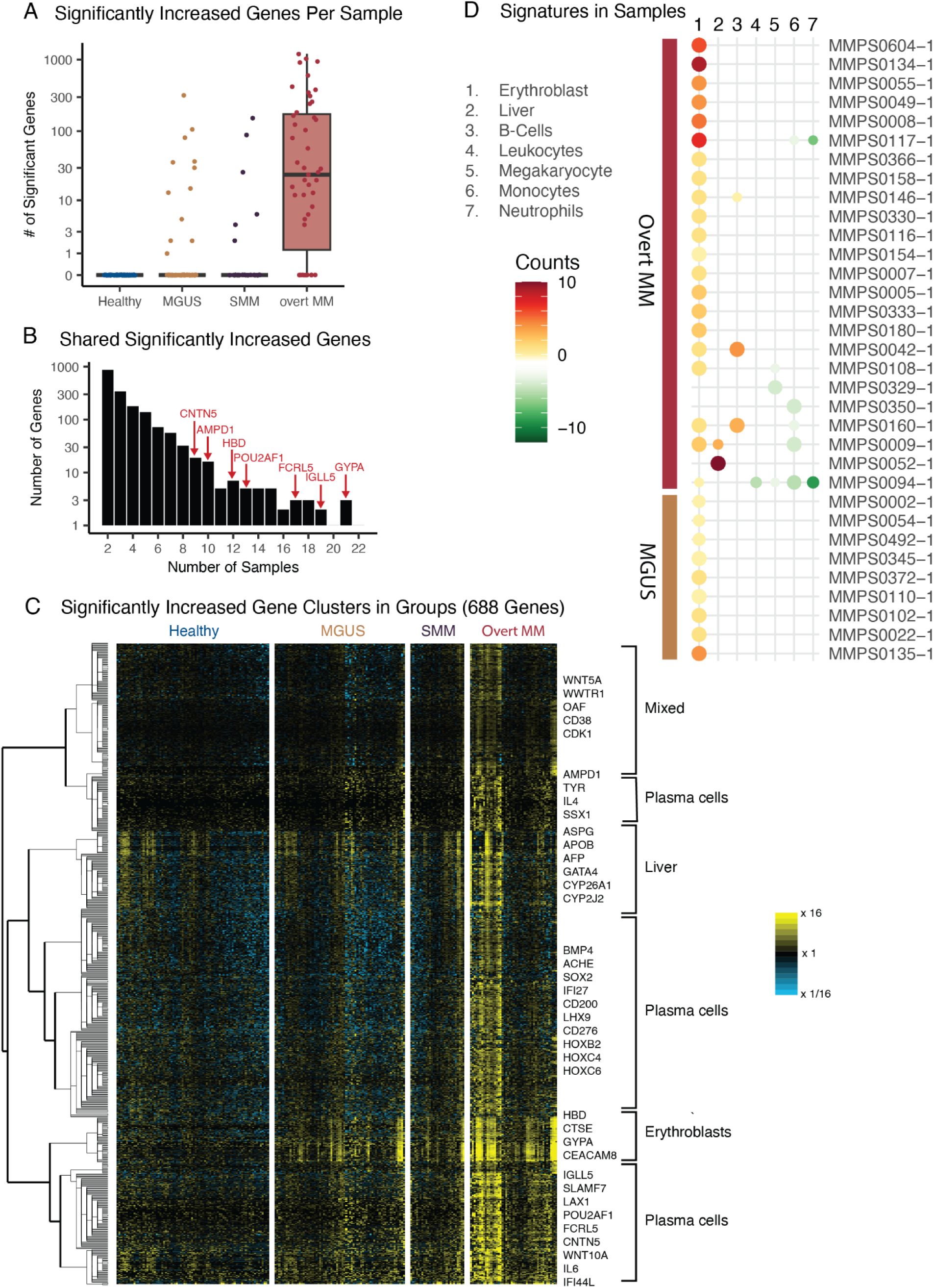
Cell-type–Cell-type-specific signal changes across the cohort **A. Differential Gene Counts by Group** – Boxplot showing the number of genes with significantly elevated levels (vs. healthy) per sample across clinical groups: Healthy, MGUS, SMM, and Overt MM. Each point represents an individual sample. **B. Recurrently Altered Genes in Overt MM** – Barplot depicting the number of genes identified as significantly elevated in an increasing number of overt MM samples. Red-labeled genes are among the most recurrently altered and include known or suspected MM-related genes (e.g., *HBD*, *FCRL5*, *GYPA*). **C. Heatmap of Differentially Represented Genes** – Heatmap of cfChIP-seq levels of 688 RefSeq genes identified as significantly elevate in ≥3 overt MM samples. Samples are grouped by clinical stage: Healthy, MGUS, SMM, and Overt MM. The color scale represents log fold-change from the mean levels in healthy samples (yellow = high, blue = low). Gene clusters are annotated based on enrichment for specific cell types such as plasma cells, erythroblasts, and liver. **D. Cell-Type Signature Enrichment in MM Samples** – Dot plot showing enrichment of cell-type–specific gene signatures across MGUS, SMM, and overt MM samples. Dot size reflects significance (q-value), and color indicates the magnitude and direction of change from healthy controls (red = increased, green = decreased). Plasma cell and erythroblast signatures dominate in advanced stages.

**Figure S3:**
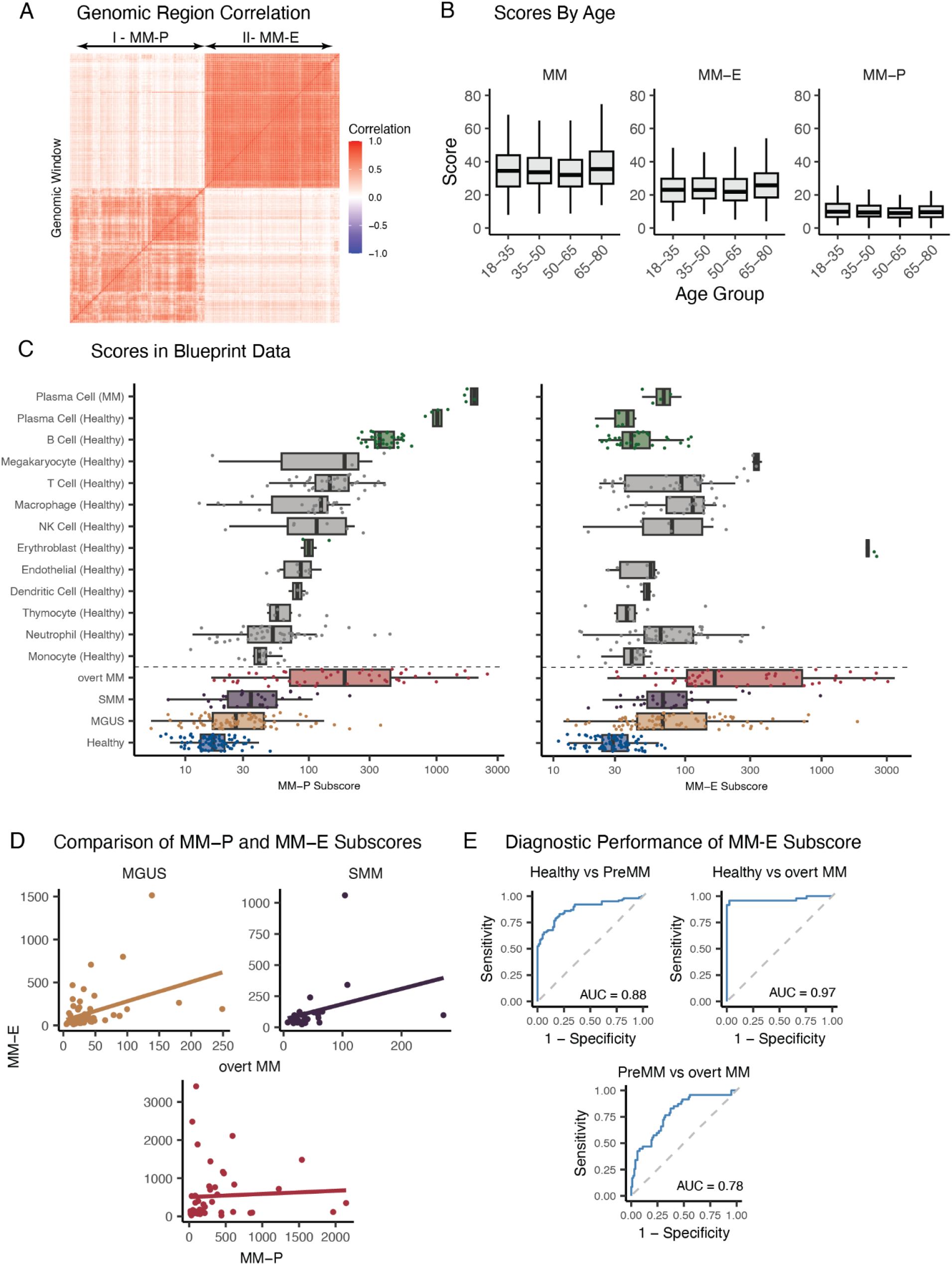
Characterization of MM-P and MM-E Subscores Across Samples **A. Gene Correlation Heatmap** – Heatmap of pairwise correlations between genes included in the MM-E and MM-P subscores. Distinct clustering of MM-E and MM-P genes indicates minimal overlap and distinct biological signals. **B. MM Scores Across Age Groups** – Boxplots showing MM score, MM-P subscore, and MM-E subscore across five age groups in healthy individuals. All scores appear relatively stable across age, suggesting limited confounding by age in score interpretation. **C. Distribution of MM-P and MM-E Subscores in Reference Cell Types** – Boxplots showing MM-P (left) and MM-E (right) subscores across Blueprint reference cell types and study samples. Plasma cells from MM patients show high MM-P scores, while erythroblasts show elevated MM-E scores. **D. Relationship Between MM-P and MM-E Subscores** – Scatter plots showing correlation between MM-P and MM-E subscores within MGUS, SMM, and overt MM groups. A modest positive correlation is observed in all groups, with increasing spread in overt MM samples. **E. ROC Curves for MM-E Subscore** – ROC curves assessing the ability of the MM-E subscore to discriminate between clinical stages. MM-E subscore distinguishes healthy from pre-MM (MGUS/SMM) samples (AUC = 0.88), overt MM from healthy (AUC = 0.97), and pre-MM from overt MM (AUC = 0.78).

**Figure S4:**
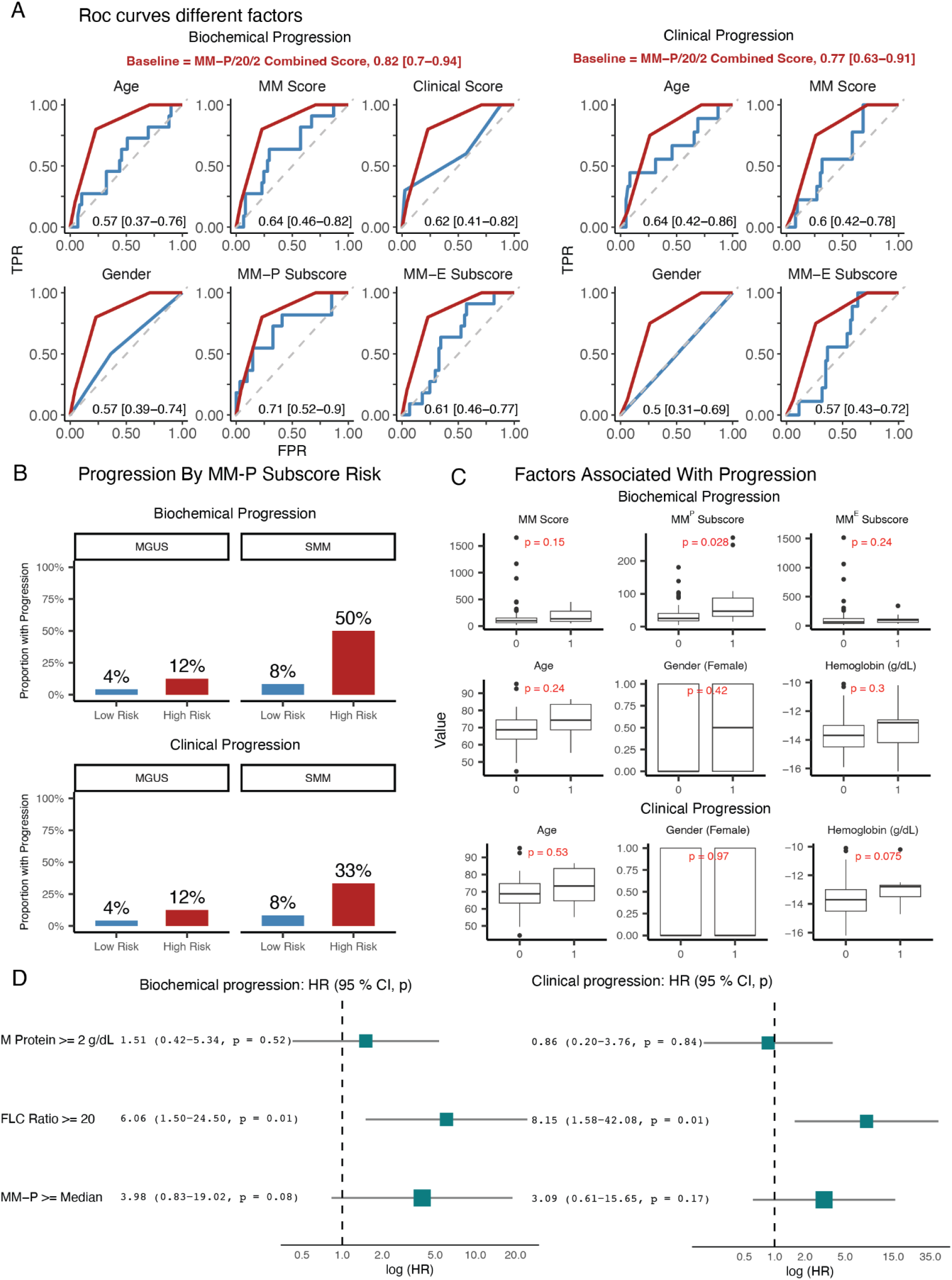
MM-P subscore and combined score enhance risk stratification across progression types **A. Progression Rates by MM-P Risk Group Across MGUS and SMM –** Bar plots showing the proportion of individuals who progressed (y-axis) within each MM-P risk group (x-axis: Low Risk vs. High Risk) for MGUS and SMM cohorts. In both clinical groups, a higher proportion of individuals progressed among those classified as High Risk, indicating that MM-P stratification provides predictive value beyond the clinical stage. Notably, progression cannot be determined solely by MGUS vs. SMM classification. **B. Comparison of Clinical Covariates by Progression Status** – Boxplots comparing MM Score, Age, Gender, and Hemoglobin levels between non-progressors (0) and progressors (1) for biochemical (top row) and clinical (bottom row) progression. Only the MM Score showed a nominal association with biochemical progression (p = 0.026). **C. ROC Analysis of Predictive Features** – ROC curves comparing the predictive power (AUC with 95% CI) of various features for biochemical (left) and clinical (right) progression. The MM-P/20/2 combined score achieved the highest AUCs for both progression types (0.82 and 0.77, respectively), outperforming individual features such as Age, Gender, MM Score, and MM-P/MM-E subscores. **D. Forest Plots of Cox Regression Hazard Ratios** – Forest plots showing hazard ratios (HRs) with 95% confidence intervals and p-values from Cox regression models for biochemical (left) and clinical (right) progression.

## Notes

### Author Declarations

IRB of Hadassah Medical Center gave ethical approval for this work

## References

1. Rajkumar, S. V. Multiple myeloma: 2020 update on diagnosis, risk-stratification and management. Am. J. Hematol. 95, 548–567 (2020).

2. Kyle, R. A. et al. Clinical course and prognosis of smoldering (asymptomatic) multiple myeloma. N. Engl. J. Med. 356, 2582–2590 (2007).

3. Kyle, R. A. & Rajkumar, S. V. Monoclonal gammopathy of undetermined significance and smouldering multiple myeloma: emphasis on risk factors for progression. Br. J. Haematol. 139, 730–743 (2007).

4. Malard, F. et al. Multiple myeloma. Nature Reviews Disease Primers 10, 1–21 (2024).

5. Rajkumar, S. V. et al. Serum free light chain ratio is an independent risk factor for progression in monoclonal gammopathy of undetermined significance. Blood 106, 812–817 (2005).

6. Rajkumar, S. V. et al. International Myeloma Working Group updated criteria for the diagnosis of multiple myeloma. Lancet Oncol. 15, e538–48 (2014).

7. Mateos, M.-V. et al. International Myeloma Working Group risk stratification model for smoldering multiple myeloma (SMM). Blood Cancer J. 10, 102 (2020).

8. Mithraprabhu, S., Chen, M., Savvidou, I., Reale, A. & Spencer, A. Liquid biopsy: an evolving paradigm for the biological characterisation of plasma cell disorders. Leukemia 35, 2771–2783 (2021).

9. Ferreira, B. et al. Liquid biopsies for multiple myeloma in a time of precision medicine. J Mol Med (Berl) 98, 513–525 (2020).

10. Manier, S. et al. Whole-exome sequencing of cell-free DNA and circulating tumor cells in multiple myeloma. Nature Communications 9, 1–11 (2018).

11. Sadeh, R. et al. ChIP-seq of plasma cell-free nucleosomes identifies gene expression programs of the cells of origin. Nature Biotechnology 39, 586–598 (2021).

12. White, B. S. et al. A multiple myeloma-specific capture sequencing platform discovers novel translocations and frequent, risk-associated point mutations in IGLL5. Blood Cancer J. 8, 35 (2018).

13. Mapping the High-Risk Multiple Myeloma Cell Surface Proteome Identifies T-Cell Inhibitory Receptors for Immune Targeting. Blood 138, 265 (2021).

14. Yu, Z. et al. Fc receptor-like 5 (FCRL5)-directed CAR-T cells exhibit antitumor activity against multiple myeloma. Signal Transduct. Target. Ther. 9, 16 (2024).

15. De Matos Simoes, R. et al. POU2AF1 as a master regulator of oncogenic transcription factor networks in myeloma. Blood 136, 18–19 (2020).

16. Kuleshov, M. V. et al. Enrichr: a comprehensive gene set enrichment analysis web server 2016 update. Nucleic Acids Res. 44, W90–7 (2016).

17. Adams, D. et al. BLUEPRINT to decode the epigenetic signature written in blood. Nat. Biotechnol. 30, 224–226 (2012).

18. Roadmap Epigenomics Consortium et al. Integrative analysis of 111 reference human epigenomes. Nature 518, 317–330 (2015).

19. Benjamini, Y. & Hochberg, Y. Controlling the false discovery rate: a practical and powerful approach to multiple testing. Journal of the royal statistical society series b-methodological 57, 289–300 (1995).

20. Therneau, T. M. Survival Analysis [R package survival version 3.8-3]. (2024).

21. Drawing Survival Curves using ‘ggplot2’ [R package survminer version 0.5.0]. (2024).

22. Fox-Fisher, I., Weinstein, O., Piyanzin, S. et al. Epigenetic liquid biopsy predicts progression to multiple myeloma. Unpublished manuscript (2025).

